# Impact and cost of scaling up TB screening and diagnostics in Asia’s ten high-burden countries: a modelling analysis

**DOI:** 10.64898/2026.04.16.26351072

**Authors:** Sandip Mandal, Kirankumar Rade, Amandeep Singh, Sreenivas A. Nair, Suvanand Sahu

## Abstract

**Background:** Tuberculosis (TB) remains a critical public health challenge, with two-thirds of the global TB burden in ten Asian countries. Social vulnerabilities, comorbidities, health inequity, multi-dimensional poverty, malnutrition, and barriers to healthcare access continue to fuel TB epidemic. Inability to detect asymptomatic and sub-clinical TB, combined with passive approach in service delivery and overreliance on smear microscopy, leads to delayed diagnosis, a substantial burden of undetected cases, and continuing TB transmission in the communities. In such a context, the introduction and scale-up of active case-finding approaches - including community-based TB screening using highly sensitive screening tools and novel rapid diagnostics - becomes a strategic priority to interrupt transmission. The growing availability of multiple screening and diagnostic options makes evidence-based decision-making increasingly complex.

**Methods:** To estimate the potential epidemiological impact and cost implications of scaling up TB diagnostics and community-based screening in ten high-burden Asian countries, we constructed a mathematical model and evaluated multiple intervention scenarios. We then assessed and compared four service delivery models: 1) digital ultraportable chest x-ray (UPCXR) & Xpert/Truenat in community, 2) digital UPCXR in community and Xpert/Truenat at health facilities, 3) digital UPCXR in community and near point of care (nPOC) at health facilities, 4) nPOC in community & Xpert/Truenat at health facilities - for total investment required and projected health benefits for their cost-effectiveness.

**Results and conclusions:** The modelling study indicated that strengthening health facility capacity (with enhanced TB screening, expanded molecular diagnostics, reduced loss to follow-up, private sector standard of care, leading to increased treatment coverage & quality of active disease treatment and reduced post-treatment relapse, scale-up of TB preventive treatment (TPT), and provision of nutritional support to 80% of TB patients and their household contacts) can significantly reduce TB incidence and mortality; however, community-wide mass screening remains essential to achieving TB elimination targets. Targeted screening of vulnerable populations demonstrated greater cost-effectiveness than untargeted screening approaches. Achieving the End TB goals will ultimately require an effective TB vaccine with high population-level coverage. AI-enabled digital UPCXR–based screening combined with Xpert/Truenat testing at the community level demonstrated maximum epidemiological impact potential, while the most cost-efficient model is Digital UPCXR in the community combined with nPOC testing at health facilities.

An investment of USD 12.7 billion over the next five years in community-level implementation of digital UPCXR and molecular diagnostics could avert an additional 9.8 million TB cases and 1.9 million deaths across ten Asian countries over a ten-year horizon

## Introduction

Tuberculosis (TB) remains a serious public health challenge and a significant barrier to achieving Universal Health Coverage and the Sustainable Development Goals [1,2]. Asia carries approximately 65% of the global TB burden, with ten Asian Development Bank (ADB) member countries - Bangladesh, Cambodia, India, Indonesia, Mongolia, Myanmar, Pakistan, Philippines, Tajikistan, and Viet Nam - together accounting for nearly half of all TB cases worldwide [2]. Millions of people in these countries remain undiagnosed or are diagnosed late, perpetuating ongoing transmission. Despite decades of national TB programs, persistently high incidence rates, delayed diagnosis, and deep socioeconomic inequities continue to impede progress.

The TB burden is disproportionately concentrated among socially vulnerable and undernourished populations, including residents of densely populated urban settlements, people in rural and remote areas, displaced and migrant populations, incarcerated individuals, and those with comorbidities such as HIV or diabetes mellitus [3–5] Heavy reliance on symptom-based screening results in many asymptomatic or subclinical cases being missed, while dependence on passive case detection and sputum microscopy contributes to diagnostic delays, treatment deferral, and sustained transmission.

Several modelling studies have evaluated the epidemiological impact of various intervention strategies and their associated costs [6–9]. Recent modelling evidence from India indicates that mass screening coupled with prompt treatment is among the fastest, most effective, and most cost-efficient strategies for achieving substantial and rapid reductions in TB incidence and mortality [10]. Mass screening operates through multiple complementary mechanisms: it enables identification of a large proportion of infectious TB cases that remain outside the formal healthcare system and facilitates the early detection of asymptomatic or subclinical disease, which may play a significant role in sustaining TB transmission. By interrupting transmission earlier in the disease course, mass screening and treatment can substantially accelerate declines in TB incidence and mortality. Advances in diagnostics, digital technologies, and service delivery platforms have made population-level implementation of mass screening more feasible than previously possible. When appropriately designed and implemented, mass screening programs may also yield broader public health benefits by serving as integrated platforms for screening of other high-priority non-communicable conditions, including diabetes mellitus, hypertension, and chronic lung diseases.

These considerations collectively highlight the urgent need for a paradigm shift toward proactive, systematic screening of high-risk populations at community level. Newer technologies, including AI-assisted digital chest X-rays, molecular diagnostics, smartphone-based cough analytics, and biomarker-based tests— are proving to enable earlier and more accurate detection, even among individuals without typical symptoms [11–15]. When integrated into tailored screening algorithms, these innovations have the potential to substantially increase case detection, shorten diagnostic delays, and curb transmission.

In this study, a mathematical model was developed and analyzed to assess the epidemiological impact of various mass screening strategies across different levels of population coverage. The study evaluated how scaling up next-generation diagnostics could influence TB incidence, mortality, and the cost-effectiveness of interventions across ten high-burden Asian countries. The findings provide evidence to guide strategic expansion of advanced diagnostics and accelerate progress toward TB elimination targets.

## Methods

### Outline of the Model

A deterministic compartmental model was developed capturing the natural history of TB epidemics. The detailed model structure and governing equations are provided in the supplementary document (Fig S1 and section S1). For simplicity, the model does not explicitly account for age structure, gender-specific differences in TB burden, or distinctions between pulmonary and extrapulmonary TB. The model framework explicitly incorporates high-risk sub-groups in which TB prevalence is higher than in the general population. A schematic diagram of the model is presented in Fig 1. Individuals in the susceptible (S) population who become infected transition to the latent infection compartments (Lf, Ls), and a subset subsequently progress to active disease (Ia, Is). Individuals with active disease may seek care in either the public (Dx(pu)) or private (Dx(pr)) sector. Once diagnosed, they generally initiate treatment in the corresponding sector, leading to outcomes of cure, treatment failure, or default—each associated with distinct relapse risks (Rlo, Rhi, R). Relapse and reinfection pathways are also incorporated in the model. Although not depicted in Fig 1, self-cure and TB-related mortality are accounted for in the model. For each of the ten Asian countries, model parameters were calibrated to align with WHO estimates of incidence and mortality, national prevalence survey data on subclinical TB, and other country-specific sources.

**Fig 1.**
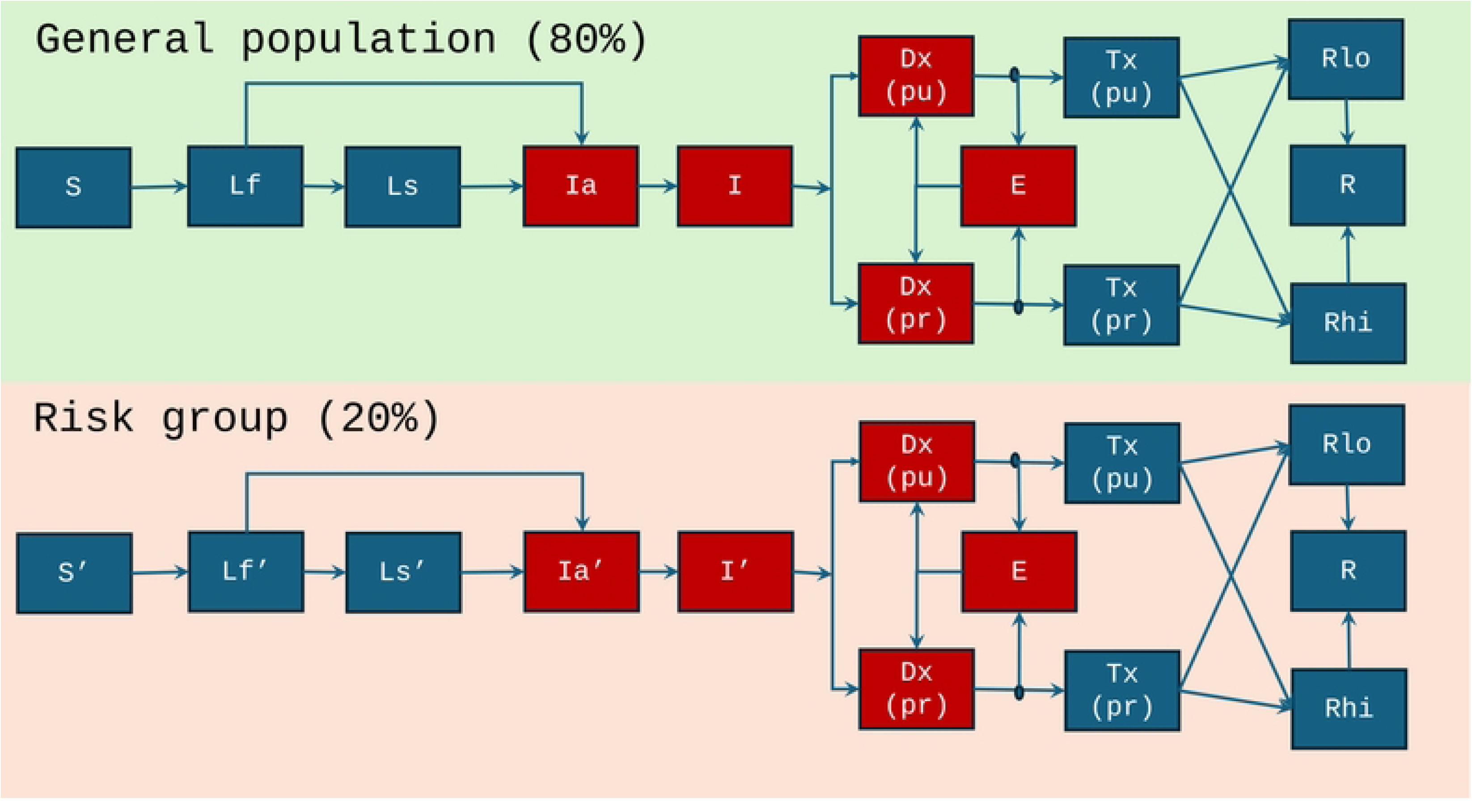
Schematic model diagram. Compartments in red show infectious states. In the top panel, green-shaded area represents general population and the bottom panel in risk group. The progression and reactivation rate to disease in risk group population is higher than the general population.

### Data and model calibration

We estimated model parameters to match the following calibration targets: WHO estimates for incidence (all forms of TB) in 2023; TB induced mortality rate in 2023; TB notifications in 2023; TB prevalence in 2020; prevalence of infection and proportion of symptomatic TB among prevalent cases. We also assumed that, in each of the mentioned countries, 20% of the population is vulnerable with a relative risk of TB twice that of the general population. The calibration targets used to fit the model to the data are summarised in Table 1, using India as an illustrative example. Natural history parameters and their estimated parameter values are provided in Table S1 of the supplementary document.

**Table 1.** Data used for model calibration (for India).

| Indicators | Calibration targets | Sources/Note |
| --- | --- | --- |
| Incidence per 100,000 population in 2023 | 195 [164 - 228] | WHO GTB Report<br>2024[2] |
| Notification per 100,000 in 2023 | 130 (+/-20%) |  |
| Mortality per 100,000 in 2023 | 23 [16 – 30] |  |
| Prevalence per 100,000in 2020 | 312 [286 – 337] | India prevalence<br>survey[18] |
| LTBI prevalence | 25% (+/- 10%) |  |
| Proportion of prevalent cases those are<br>symptomatic. | 0.5 [0.36 – 0.8] |  |
| Relative risk of TB | 2.0 (+/- 20%) | Assumption |
| Percentage of population in risk group | 20 (+/- 20%) | Assumption |

Parameter values for other countries are given in Table S2 (a - i) of the supplementary document and calibration targets are shown in Table S3.

Calibration was performed using an adaptive Bayesian Markov Chain Monte Carlo (MCMC) method [16]. Specifically, likelihood functions were constructed for each calibration target, incorporating both central estimates and associated uncertainty intervals. A posterior density was then constructed by combining these likelihood functions with uniform priors for the model parameters. Sampling from the posterior density was performed using an adaptive Metropolis algorithm [17]. After eliminating the burn-in phase and applying ‘thinning’, 250 samples were retained for simulation. For all model projections, uncertainty was quantified by defining the interval between the 2.5th and 97.5th percentiles as the ‘95% Bayesian credible interval’. The central estimate of each projection was taken using its 50th percentile (median)

### Modelling interventions

To assess the epidemiological impact of expanding TB control activities, seven intervention scenarios were modelled, each cumulatively building on the preceding one. The baseline scenario assumed no additional interventions beyond ongoing routine programme activities. The second scenario - the Health Facilities Strengthening scenario - incorporates improvements across the TB care continuum. This included, for public sector health facilities, enhanced TB screening, expanded use of Xpert/Truenat, and reduced initial loss to follow-up (LTFU). It also strengthens private sector engagement, ensuring that individuals receive the same standard of care as in the public sector. Additionally, the scenario incorporates reductions in progression to active disease and post-treatment relapse through increased treatment coverage and quality, scale-up of TPT following active evaluation of household contacts, and provision of nutritional support to 80% of TB patients and their household contacts.

Population-level screening strategies were then added in successive steps. The third scenario included annual screening of an estimated 20% of the population identified as clinically or socially vulnerable - including PLHIV, diabetics, smokers, alcohol users, undernourished individuals, and those living in urban slums, tribal communities or other congregate settings. In the fourth scenario, screening coverage expanded to 40% of the population comprising the vulnerable subgroup plus an additional 20% of the general population), and finally, in the fifth scenario, coverage was expanded to 60%, comprising the vulnerable subgroup plus 40% of the general population). Screening algorithms incorporated symptom assessment together with digital chest X-ray or other novel screening tools, followed by confirmatory molecular testing and drug susceptibility testing (DST) when indicated.

Two prevention-focused scenarios were also evaluated. The first assumed 50% of adults received a Prevention-of-Disease (PoD) vaccine with 50% efficacy in reducing progression from infection to active TB. The final and most optimistic scenario combined PoD vaccination with a Prevention-of-Relapse (PoR) effect (50% efficacy), along with community-based nutritional support for 30% of undernourished individuals to reduce disease progression.

All interventions are assumed to be scaled up over a three-year period beginning in 2026, with the exception of vaccination, which is assumed to start in 2029.

### Service Delivery Models

Service-delivery models for community-based population screening were defined based on technologies and tools that are currently available and have been demonstrated to be operationally feasible. These included digital ultraportable chest X-ray (UPCXR) with artificial intelligence (AI) - based automated reading, along with Xpert/Truenat tests incorporated into the screening & diagnostic algorithms. In addition, newer low-cost near-point-of-care (nPOC) technologies were considered. Based on the combination of screening and testing locations and methods, four service delivery models were prioritized:

1. **Model 1: Digital UPCXR & Xpert/Truenat in Community:** Both screening and confirmatory testing (Xpert/Truenat) are conducted within the community by the mobile teams, minimizing delays between screening and diagnosis.
2. **Model 2: Digital UPCXR in Community & Xpert/Truenat at Health Facilities:** The target population is screened for TB symptoms and chest abnormalities using digital UPCXR operated by mobile teams. Sputum samples from eligible individuals (those who are symptomatic and/or have chest x-ray abnormalities) are tested with mWRD at health facilities.
3. **Model 3: Digital UPCXR in Community & nPOC in Health Facilities:** The target population is screened in the community with digital UPCXR, followed by nPOC testing of eligible sputum samples at health facilities for confirmation and treatment initiation.
4. **nPOC in Community & Xpert/Truenat at Health Facilities:** The target population is screened in the community through sample collection and testing using a novel nPOC test. Individuals testing positive undergo confirmatory testing and treatment initiation at the health facilities.

### Requirement Assessment

For each technology and service delivery model, requirements estimated through the following steps: Based on the country specific national TB prevalence surveys and reported program experiences, the following were estimated for the population aged 15 years and older -, 1) the proportion eligible for sputum collection based on CXR findings, 2) the proportion eligible based on symptoms alone, and 3) the proportion eligible based on a combination of symptoms and CXR findings [19] Estimates were adjusted to reflect the proportion of population ≥15 in each country. Assumptions were made regarding participation rates for screening, sputum sample collection rates (among eligible individuals), actual testing rates, the proportion of valid test results and the sensitivity of the first test in the algorithm. These were adjusted for screening outcomes within each service delivery model including attrition and sample loss during transport to health facility. Current eligibility rates for sputum testing and TB positivity rates were derived from ongoing community-based TB screening projects. (Table S4 - S5, Supplementary document). Based on this proportion of population screened, the number of sputum tests required (including confirmatory tests) and the number of TB cases diagnosed were estimated for each service delivery model. Similarly, DST requirements were estimated using current Drug resistance TB data reported by WHO in the 2025 Global TB Report [2]. The details are given in Table S6 in the supplementary document. Currently available nucleic acid amplification testing (NAAT) capacity was assessed based on current utilization rates per machine, and aggregate national capacity. Spare (unutilized) NAAT capacity was calculated and was removed from the gross testing requirement for community-based TB screening, under the assumption that existing capacity will be fully used before additional machines are procured.

### Outline of Costing and Forecasting

For community-based TB screening, it was assumed that dedicated full time field teams would be recruited and equipped with the necessary tools including the UPCXR, Xpert/Truenat machines, nPOC devices, together with associated consumables including cartridges as well as operational costs including transportation and overheads.

Unit costs for materials - including UPCXR machines, NAAT machines, nPOC devices and consumables were sourced from the Global Drug Facility (GDF) catalogue [20,21]. Data on the existing available number of machines in the country was obtained from the Airborne Infection Defence Platform (AIDP) report on landscape assessment of ASEAN member countries for Pandemic Preparedness and TB Response and WHO Global TB Database [22]. HR costs were informed by experience from ongoing community-based TB screening projects, as were operational costs including transport, field processes and overheads. Details of costing assumptions are given in Table S7 in the supplementary document. Most of the costing data was available for the time period between 2018-2023 [23]. To forecast country-specific changes in costs of items, the World Bank Deflator was used. WB Deflator trends were plotted for each country separately for the period 2010-2024, and a linear trend was used to forecast country-specific costs between 2026 and 2030, enabling estimation of the additional investment required for each service delivery model and coverage scenario. Details are given in Table S8 in the supplementary document [24]. The total investment cost per country was calculated as the product of the additional required units and the forecasted unit cost.

### Cost-effectiveness Analysis of Service Delivery Models

For community-based TB screening, coverage was scaled up in three steps: first targeting the initial 20% as the vulnerable population, then expanding to include an additional 20% of the general population, and subsequently to 40% of the general population, resulting in a total of 60% coverage. Epidemiological model outputs were calibrated for each of the four service delivery options using the sensitivity of TB testing within the respective diagnostic algorithm (Table S4, supplementary document). Using these differential sensitivities for each service delivery model, alongside the corresponding epidemiological impact estimates for TB incidence and mortality, additional TB cases and deaths averted were calculated for the period 2026 – 2030, with projections extended to 2050 to capture the long-term impact of the interventions. Costs were estimated and investment budgets were prepared for the five-year period from 2026 to 2030.

Additional TB cases and deaths averted were estimated for each service delivery model and coverage level, calculated as incremental gains corresponding to each stepwise increase in community screening coverage. Incremental investment costs were calculated as described in the preceding section. Cost per TB case averted and cost per TB death averted were then estimated for each country across the four service delivery models at three coverage scenarios: (i) 20% population screening (vvulnerable population only), (ii) 40% population screening (20% vulnerable, 20% general population), and (iii) 60% population screening (20% vvulnerable, 40% general population). The incremental Cost-Effectiveness Ratio (ICER) was then calculated relative to the baseline scenario (current program investment) across 12 scenarios (4 service delivery models x 3 population coverage levels).

## Results

### Calibration Results

Calibration results for the ten countries are presented in Fig 2, illustrating the agreement between model outputs and country-specific observed epidemiological data for each of the selected settings.

**Fig 2.**
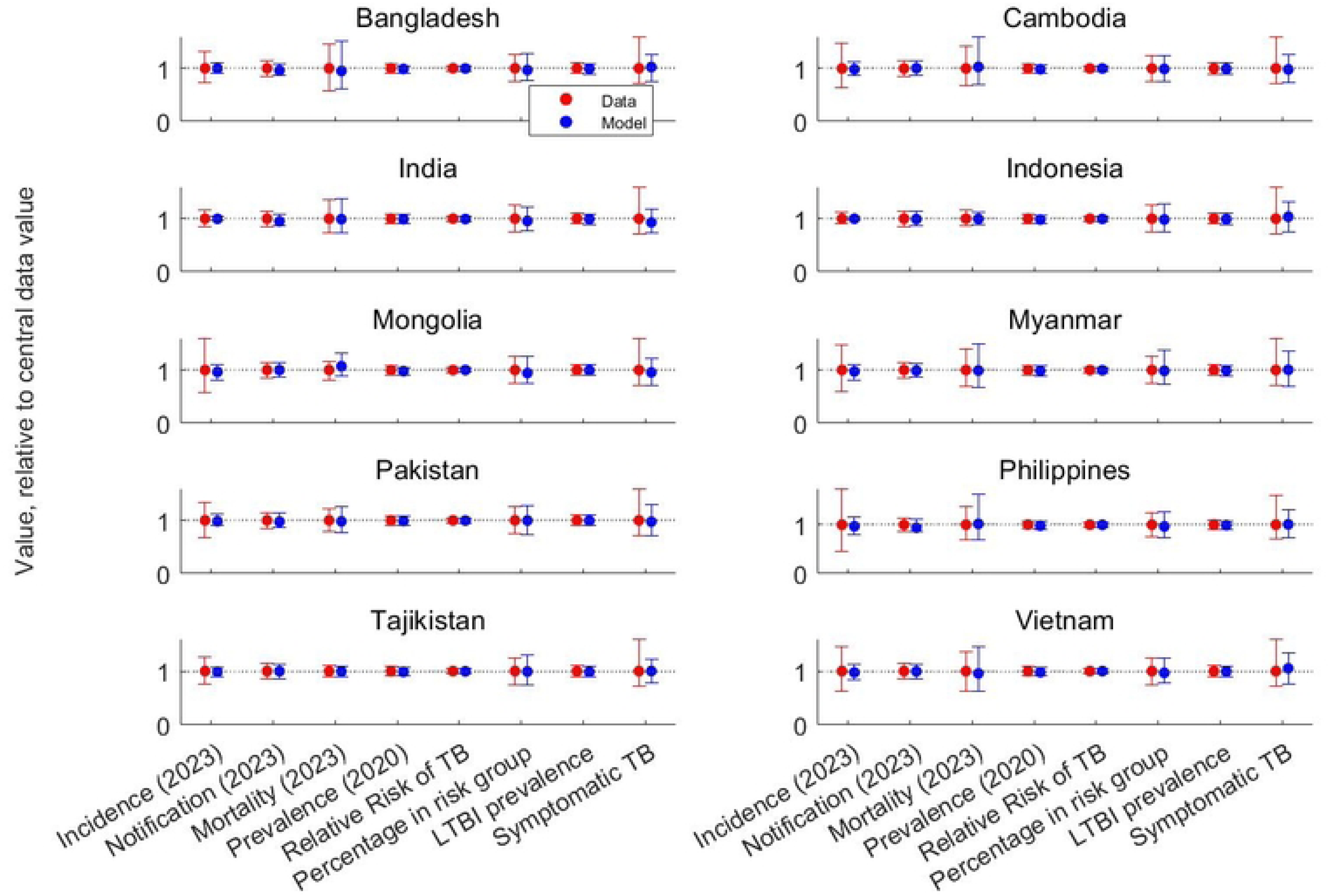
Results of model calibration. All outputs are normalized to their respective central values to enable comparison across indicators with different scales. Dots represent central estimates, while error bars show 95% uncertainty intervals. Red dots are the observed data and blue dots indicate model-outputs

### Epidemiological Impact and Cost-Effectiveness of Interventions Across Service Delivery Models

Given the volume of results across ten countries and multiple intervention scenarios, India is used as an illustrative example to describe the projected epidemiological impact of each intervention in detail. India was selected as the primary example given its scale, the availability of robust calibration data, and its substantial contribution to the regional TB burden. Projected TB incidence rates (per 100,000 population) and corresponding TB deaths for India under different intervention scenarios are presented in Fig 3, with aggregate results across all ten countries subsequently presented in Tables 2 and 3.

**Fig 3.**
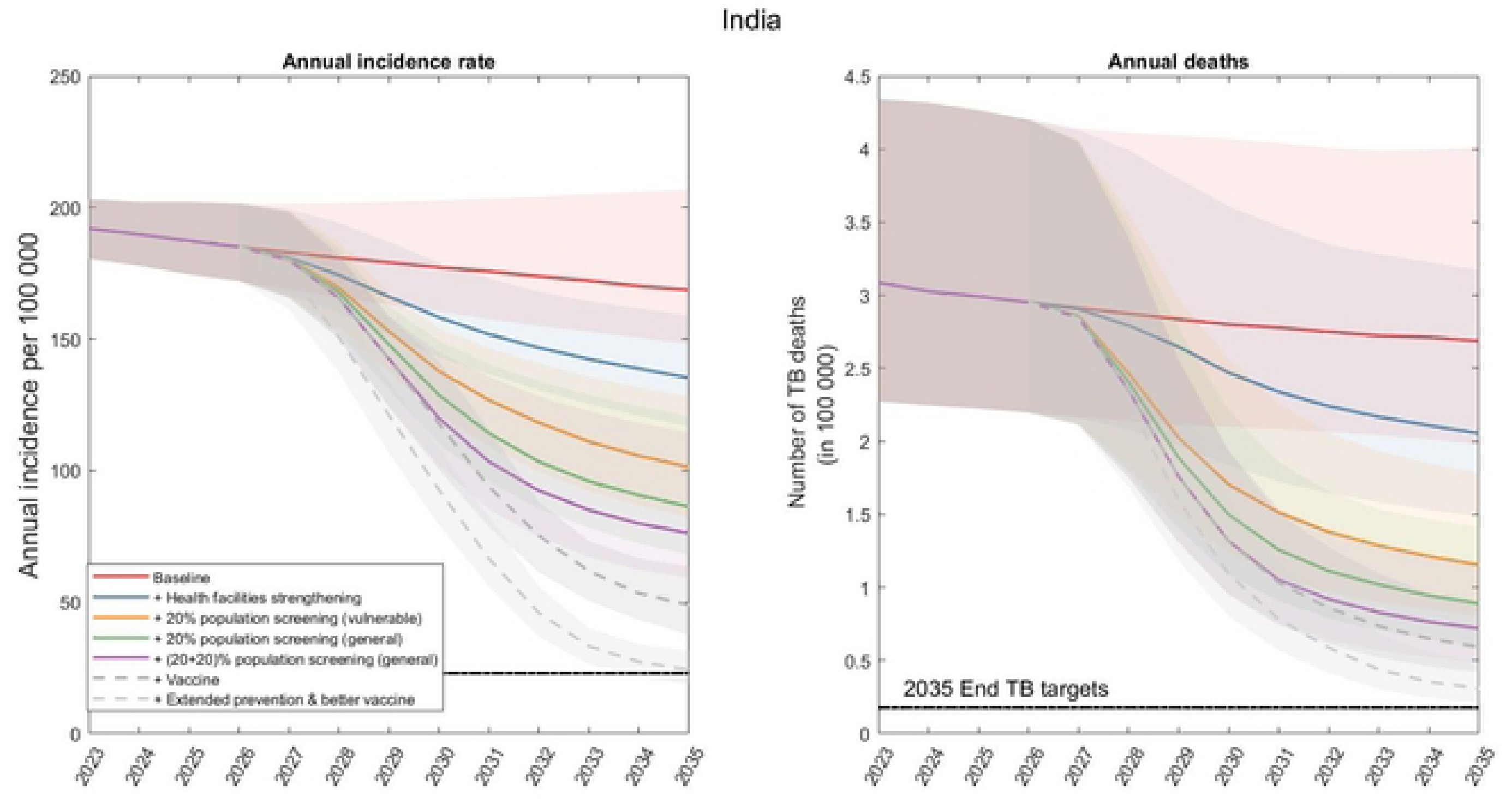
Projected incidence rate (per 100,000 population) and total TB deaths under different intervention scenarios in India. Solid or dotted line represents the central estimates, and the shaded area denotes the 95% Bayesian credible intervals.

**Table 2.** Impact of interventions with current tools in idealized conditions.

|  | Percentage reduction of incidence rate in 2035 relative to 2015 |  |  |  |  |
| --- | --- | --- | --- | --- | --- |
| Country | Strengthening health facilities | +Targeted screening of high-risk populations | +20% general population screening | +20% general population screening | +vaccine |
| Bangladesh | 27[15-55] | 42[25-67] | 49[29-72] | 54[32-78] | 81[76-88] |
| Cambodia | 19[11-41] | 33[20-63] | 39[23-71] | 44[26-76] | 80[77-86] |
| India | 44 [27 - 61] | 58 [44 – 72] | 64[47 – 76] | 68[49-79] | 80[74-85] |
| Indonesia | 15[9-32] | 28[17-58] | 35[21-67] | 40[24-74] | 81[77-87] |
| Mongolia | 12[8-27] | 28[18-58] | 37[22-71] | 43[27-80] | 82[78-88] |
| Myanmar | 28[16 – 49] | 48 [29 – 69] | 57[34 – 78] | 63[37 – 82] | 91[89 – 94] |
| Pakistan | 21[12-39] | 36[22-60] | 41[25-71] | 46[28-77] | 81[77-86] |
| Philippines | 10[7-23] | 23[12-52] | 31[16-65] | 37[19-75] | 81[78-88] |
| Tajikistan | 29[17-45] | 46[26-65] | 53[30-73] | 58[33-77] | 80[77-86] |
| Vietnam | 22[12-41] | 35[21-59] | 41[25-68] | 45[28-75] | 80[76-84] |
| Percentage reduction of TB deaths in number in 2035 relative to 2015 |  |  |  |  |  |
| Bangladesh | 28[16-59] | 58[45-78] | 67[55-83] | 73[62-87] | 81[74-90] |
| Cambodia | 21[12-43] | 50[35-71] | 58[43-79] | 64[50-84] | 75[68-87] |
| India | 49[31-66] | 71[59-81] | 78[66-86] | 82[71-88] | 85[79-90] |
| Indonesia | 17[10-36] | 49[38-69] | 59[49-79] | 67[56-85] | 78[72-89] |
| Mongolia | 14[9-29] | 48[38-66] | 60[50-80] | 70[58-87] | 79[72-90] |
| Myanmar | 32[17 – 54] | 64[48 – 79] | 74[58 – 86] | 80[66 – 90] | 86[77 – 92] |
| Pakistan | 23[14-42] | 55[43-71] | 64[51-81] | 69[58-87] | 79[73-89] |
| Philippines | 12[8-25] | 46[35-66] | 58[46-78] | 66[53-84] | 78[70-89] |
| Tajikistan | 31[18-47] | 60[44-74] | 70[54-82] | 76[60-87] | 82[73-88] |
| Vietnam | 24[13-44] | 55[41-71] | 64[51-79] | 71[58-86] | 88[82-93] |

**Table 3.** TB cases and deaths averted under idealized conditions using existing tools, by intervention scenario and country, 2026–2035.

|  | Cases averted between 2026 to 2035 in million |  |  |  |  |
| --- | --- | --- | --- | --- | --- |
| Country | Strengthenin<br>g health<br>facilities | +Targeted<br>screening of<br>high-risk<br>populations | +20% general<br>population<br>screening | +20%<br>general<br>population<br>screening | +Vaccine |
| Bangladesh | 0.27[0.18 –<br>0.43] | 0.63[0.27 –<br>1.12] | 0.79 [0.34 –<br>1.38] | 0.92 [0.40<br>– 1.59] | 1.40 [1.12 –<br>1.82] |
| Cambodia | 0.05 [0.03 –<br>0.09] | 0.09 [0.04 –<br>0.21] | 0.11 [0.05 –<br>0.25] | 0.13 [0.06-0.28] | 0.22 [0.17 –<br>0.32] |
| India | 2.93 [1.85-4.40] | 6.05 [3.13-8.75] | 7.36 [3.66-10.69] | 8.42 [4.03<br>– 12.02] | 9.94 [7.78 –<br>12.92] |
| Indonesia | 0.64 [0.48 –<br>1.22] | 1.42[0.73-3.04] | 1.78[0.89 –<br>3.79] | 2.09 [1.05<br>– 4.44] | 3.93 [3.34 –<br>5.12] |
| Mongolia | 0.012[0.008<br>– 0.022] | 0.028[0.015 –<br>0.054] | 0.035[0.019 –<br>0.068] | 0.042[0.02<br>2 – 0.081] | 0.068[0.053<br>– 0.089] |
| Myanmar | 0.29 [0.16 –<br>0.59] | 0.63 [0.25 –<br>1.16] | 0.79 [0.31 –<br>1.37] | 0.91 [0.36<br>– 1.57] | 1.17 [0.86 –<br>1.65] |
| Pakistan | 0.49[0.32 –<br>0.98] | 1.00[0.47 –<br>2.32] | 1.24[0.57 –<br>2.88] | 1.47 [0.64<br>– 3.35] | 2.60 [2.02 –<br>3.77] |
| Philippines | 0.43[0.29 –<br>0.78] | 1.00[0.45 –<br>2.11] | 1.29[0.57 –<br>2.78] | 1.56[0.68 –<br>3.31] | 2.82[2.13 –<br>3.80] |
| Tajikistan | 0.07[0.04 –<br>0.10] | 0.16[0.08 –<br>0.24] | 0.21[0.09 –<br>0.31] | 0.24[0.11-0.35] | 0.32[0.26 –<br>0.39] |
| Vietnam | 0.11[0.07 –<br>0.21] | 0.24[0.12 –<br>0.47] | 0.30[0.15 –<br>0.60] | 0.36[0.17 –<br>0.70] | 0.61[0.47 –<br>0.82] |
| Deaths averted between 2026 to 2035 in million |  |  |  |  |  |
| Bangladesh | 0.03[0.01-0.06] | 0.13[0.07 –<br>0.23] | 0.15 [0.09 –<br>0.27] | 0.17[0.10 –<br>0.30] | 0.18 [0.11 –<br>0.32] |
| Cambodia | 0.003 [0.001<br>– 0.007] | 0.011[0.006 –<br>0.020] | 0.013[0.008 –<br>0.023] | 0.015[0.00<br>9 – 0.027] | 0.016[0.010<br>– 0.028] |
| India | 0.35[0.23 –<br>0.59] | 1.02[0.67 -<br>1.54] | 1.20[0.82 –<br>1.86] | 1.32 [0.94-2.07] | 1.39[0.99 –<br>2.12] |
| Indonesia | 0.07 [0.05 –<br>0.16] | 0.35 [0.27 –<br>0.51] | 0.44 [0.34 –<br>0.60] | 0.50[0.39 –<br>0.68] | 0.55 [0.45 –<br>0.71] |
| Mongolia | 0.0003<br>[0.0002 –<br>0.0007] | 0.0014<br>[0.0010 –<br>0.0019] | 0.0018 [0.0013<br>– 0.0023] | 0.0021<br>[0.0015 –<br>0.0026] | 0.0022<br>[0.0017 –<br>0.0027] |
| Myanmar | 0.05 [0.02 –<br>0.98] | 0.15 [0.09 –<br>0.25] | 0.18 [0.11 –<br>0.29] | 0.21 [0.13<br>– 0.33] | 0.21 [0.14 –<br>0.33] |
| Pakistan | 0.039 [0.023<br>– 0.073] | 0.151[0.106 –<br>0.223] | 0.184 [0.133 –<br>0.268] | 0.214<br>[0.152 –<br>0.299] | 0.230 [0.171<br>– 0.311] |
| Philippines | 0.02[0.01 –<br>0.05] | 0.11[0.07 –<br>0.19] | 0.14 [0.09 –<br>0.22] | 0.16[0.10 –<br>0.26] | 0.17[0.12 –<br>0.28] |
| Tajikistan | 0.006[0.004<br>– 0.010] | 0.026[0.019 -<br>0.032] | 0.032[0.024 –<br>0.038] | 0.036[0.02<br>8 – 0.043] | 0.038[0.031<br>– 0.044] |
| Vietnam | 0.007 [0.004<br>– 0.014] | 0.031 [0.020<br>– 0.051] | 0.038 [0.025 –<br>0.062] | 0.044<br>[0.030 –<br>0.071] | 0.047 [0.033<br>– 0.075] |

The scenarios presented represent idealized conditions, assuming 100% effectiveness of the diagnostic algorithms; for example, that screening 20% of the vulnerable population is assumed to result in detection of all TB cases within that group. Strengthening the health system alone as described earlier with strengthened health facilities and improved TB care is projected to reduce TB incidence by 44% [27 – 61] and mortality by 49% [31 - 66] by 2035 relative to 2015 levels. The addition of community-based mass screening leads to further reductions in both incidence and mortality. Targeted screening of high-risk populations, assumed to comprise 20% of the Indian population, is projected to reduce incidence by 58% [44 - 72] and mortality by 71% [59 - 81] by 2035 relative to 2015 levels. Expanding screening to an additional 20% or 40% of the general population is projected to further reduce incidence by 64% [47 - 76] and 68% [49 - 79], respectively, with corresponding reductions in mortality of 78% [66 - 86] and 82% [71 - 88] over the same period.

Despite these improvements, none of the intervention scenarios considered is sufficient to achieve the WHO End TB incidence targets for 2035. When all of the above interventions are combined with the introduction of a vaccine with 50% efficacy against progression to active disease, initiated in 2029 and administered to 50% of the adult population, the projected reduction in incidence increases to 80% [74 - 85] relative to 2015 levels, with a corresponding reduction in mortality of 85% [79 – 90]. Furthermore, under a hypothetical scenario in which the same vaccine is additionally assumed to reduce post-treatment relapse by 50%, model projections indicate that the End TB targets for 2035 could be achieved.

The impact of interventions on TB incidence rate and mortality for all the countries is presented in Table 2. Table 3 presents TB cases and deaths averted between 2026 to 2035 under each intervention scenario.

The impact estimates presented in Tables 2 and 3 assume idealised diagnostic conditions — specifically, 100% effectiveness of the diagnostic algorithms. When test sensitivity and the specific service delivery model are accounted for, these estimates are attenuated accordingly Figs 4 and 5 present TB cases and deaths averted, respectively, across the four service delivery models at three population coverage levels.

**Fig 4.**
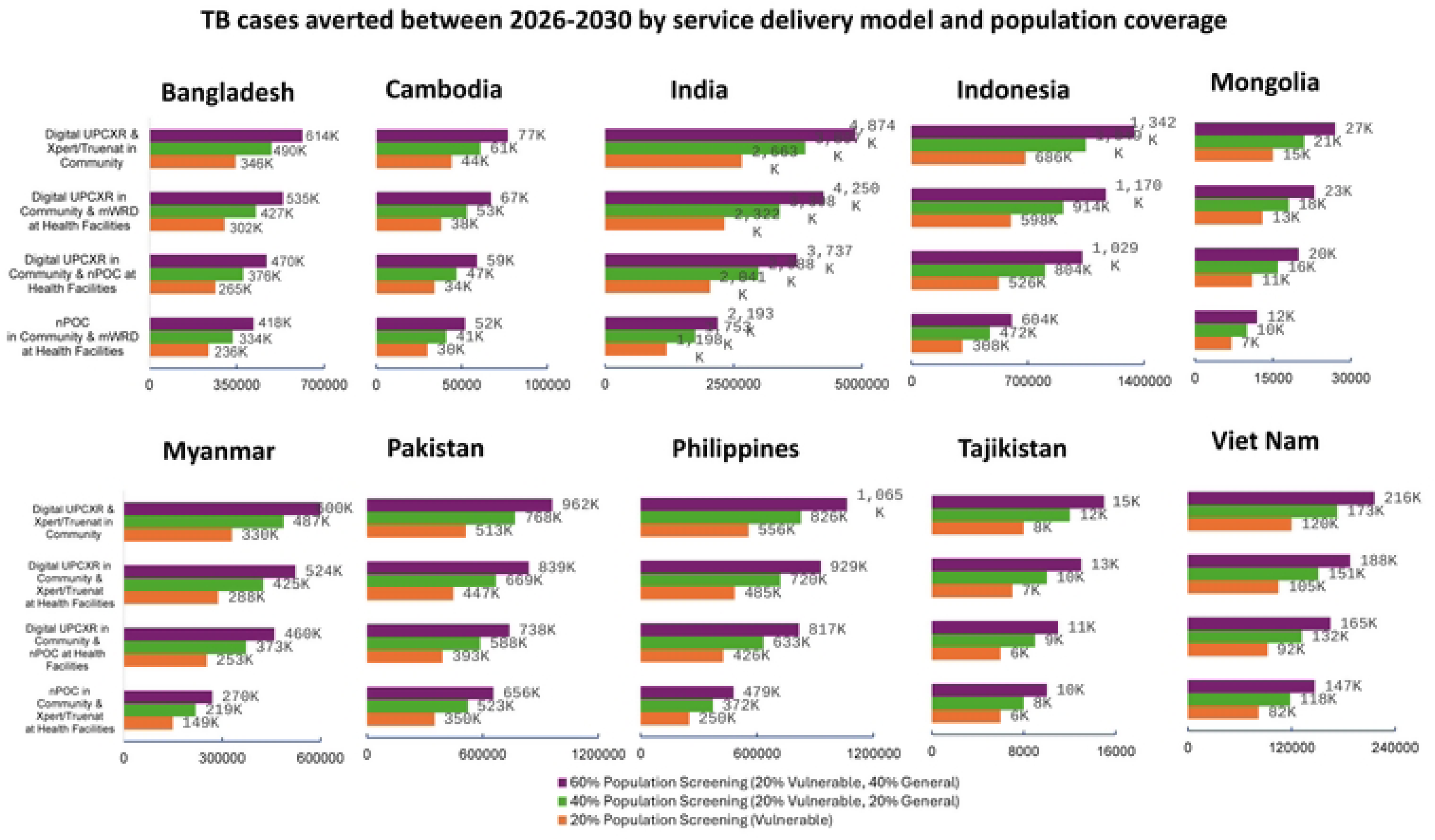
TB cases averted between 2026-2035. Estimated number of TB cases averted between 2026 and 2035, disaggregated by service delivery model and population coverage level across ten high-burden Asian countries.

**Fig 5.**
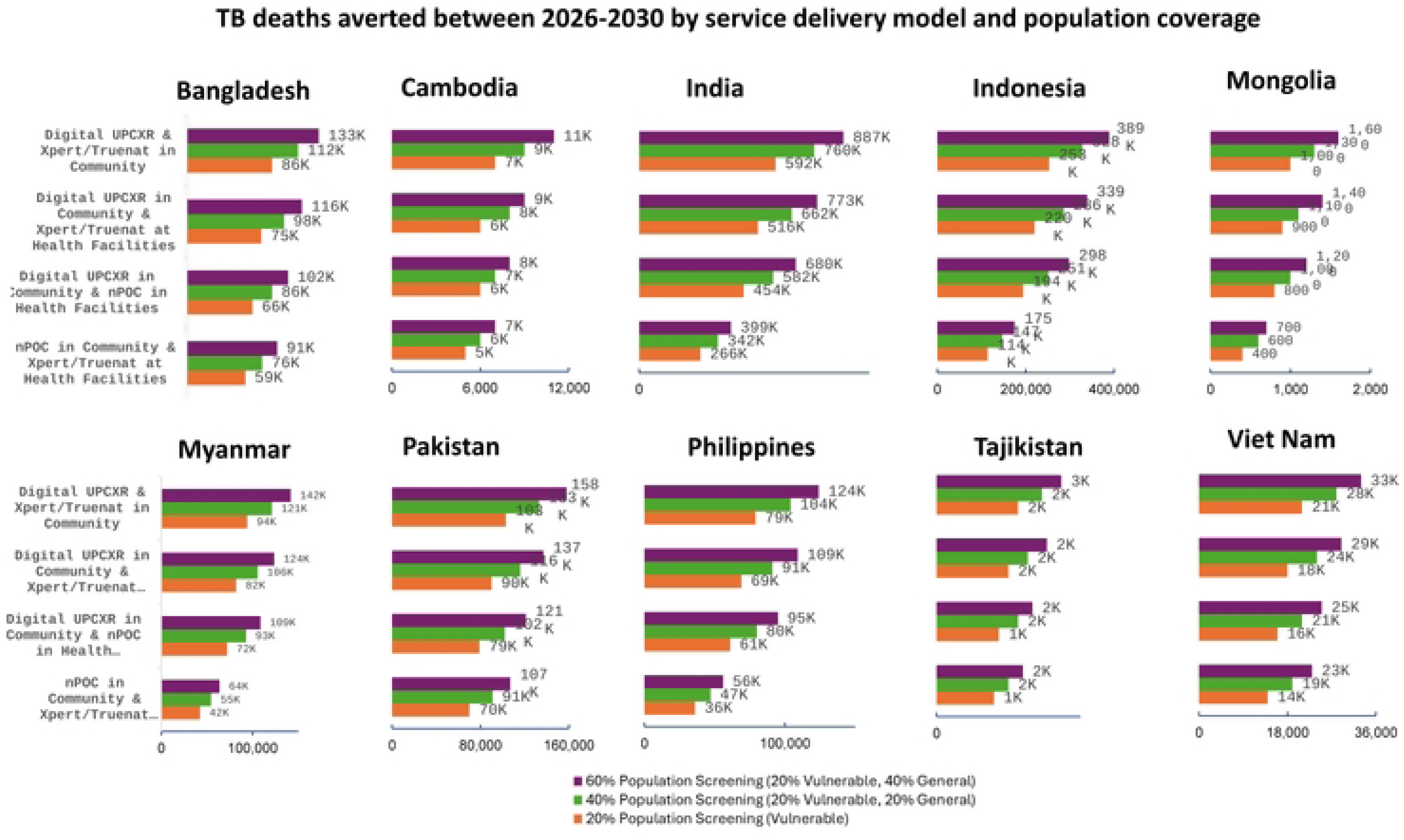
TB deaths averted between 2026-2035. Estimated number of TB deaths averted between 2026 and 2035, disaggregated by service delivery model and population coverage level across ten high-burden Asian countries.

The pattern of TB cases averted were consistent across all ten countries. Fig 4 suggests that the greatest reduction in cases was projected under Model 1 - digital UPCXR combined with Xpert/Truenat testing deployed at the community-level at 60% population coverage (Fig 4). Across all ten countries combined, an estimated 5.28 million, 7.78 million, & 9.79 million TB cases could be averted between 2026-2035, under community-based TB screening coverage of 20%, 40% and 60% population, respectively, with screening implemented over the period 2026-2030.

The largest reduction in TB deaths was projected under Model 1 - digital UPCXR combined with Xpert/Truenat testing deployed at the community level at 60% population coverage (Fig 5). Across all ten countries combined, an estimated 1.24 million, 1.60 million, and 1.88 million TB deaths could be averted during 2026–2035 under community-based screening coverage levels of 20%, 40%, and 60%, respectively, with screening implemented over the period 2026–2030. The pattern of TB deaths averted was likewise consistent across all ten countries.

Investment costs scaled approximately proportionately with population coverage across all service delivery models, suggesting limited economies of scale at higher coverage levels (Table 4).

**Table 4.** Total investment required for community-based screening scale up across ten high-burden Asian countries 2026-2030 (in USD million)

| **Service Delivery Model** | **20% population**<br>**screening**<br>**(Vulnerable)** | **40% population**<br>**screening**<br>**(20% Vulnerable,**<br>**20% General)** | **60% population**<br>**screening**<br>**(20% Vulnerable,**<br>**40% General)** |
| --- | --- | --- | --- |
| Model 1: Digital<br>ultraportable X-ray &<br>Xpert/Truenat in<br>Community | 4265 | 8483 | 12694 |
| Model 2: Digital UPCXR in<br>community &<br>Xpert/Truenat at Health<br>Facilities | 3682 | 7340 | 10982 |
| Model 3: Digital UPCXR in<br>Community & nPOC in<br>Health Facilities | 2614 | 5245 | 7815 |
| Model 4: nPOC in<br>community &<br>Xpert/Truenat at Health<br>Facilities | 5401 | 10783 | 16174 |

Among the service delivery models, Model 3 - digital UPCXR in the community combined with nPOC testing at health facilities incurred the lowest investment cost, while Model 4 - nPOC testing in the community combined with Xpert/Truenat testing at health facilities, incurred the highest. This difference is primarily driven by the volume of diagnostic tests required. In models that incorporate symptom screening and CXR as initial triage tools, only a subset of the screened population (approximately 5–15%) proceeds to confirmatory TB testing, whereas models relying on nPOC testing as the primary screening tool require a substantially larger number of tests across the screened population.

The additional investment required for each of the service delivery models across all countries, as described above, is presented in Fig 6.

**Fig 6.**
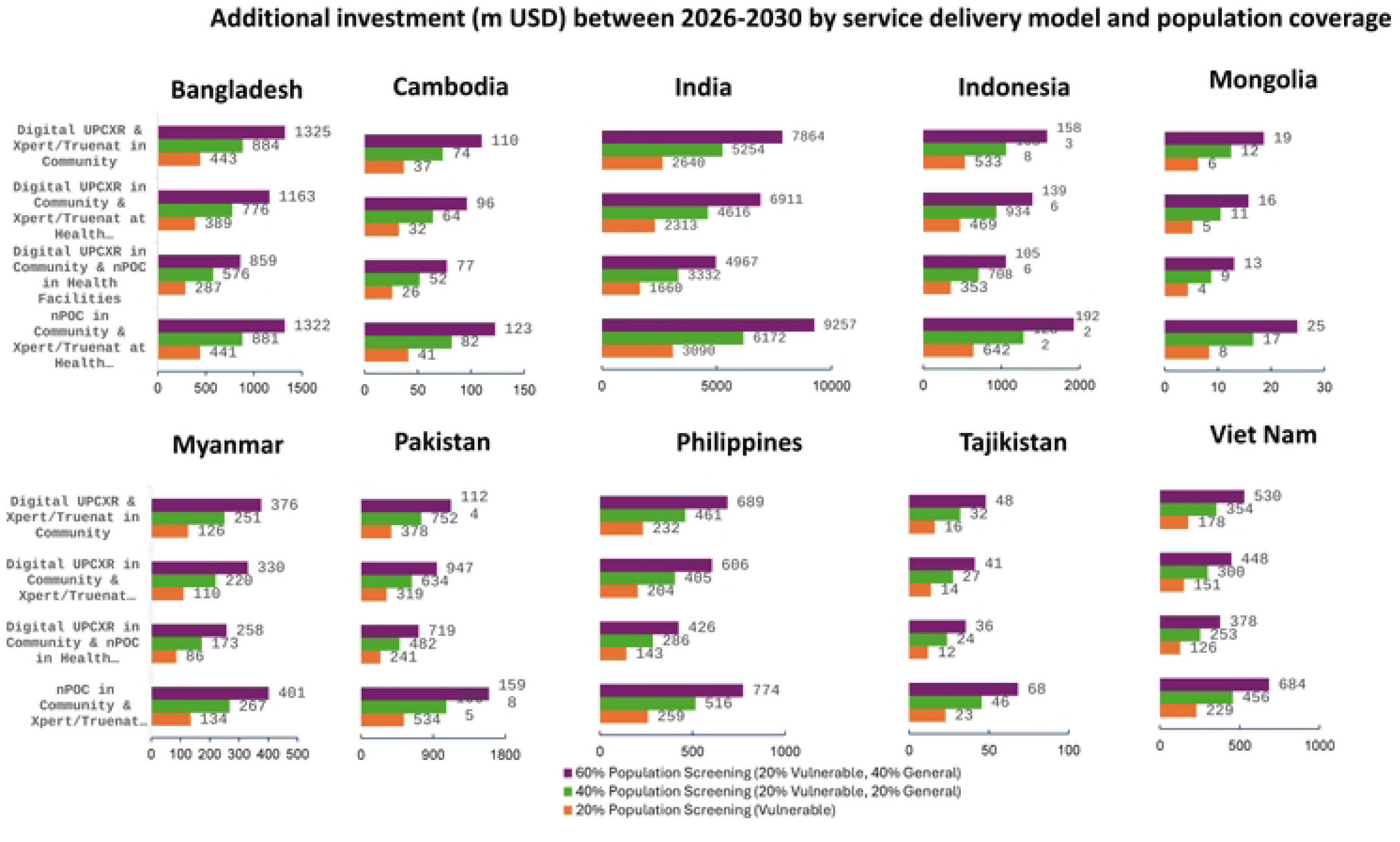
Additional investment (m USD) between 2026-2030. Estimated additional investment required, disaggregated by service delivery model and population coverage level across ten high-burden Asian countries.

**Table 5.**
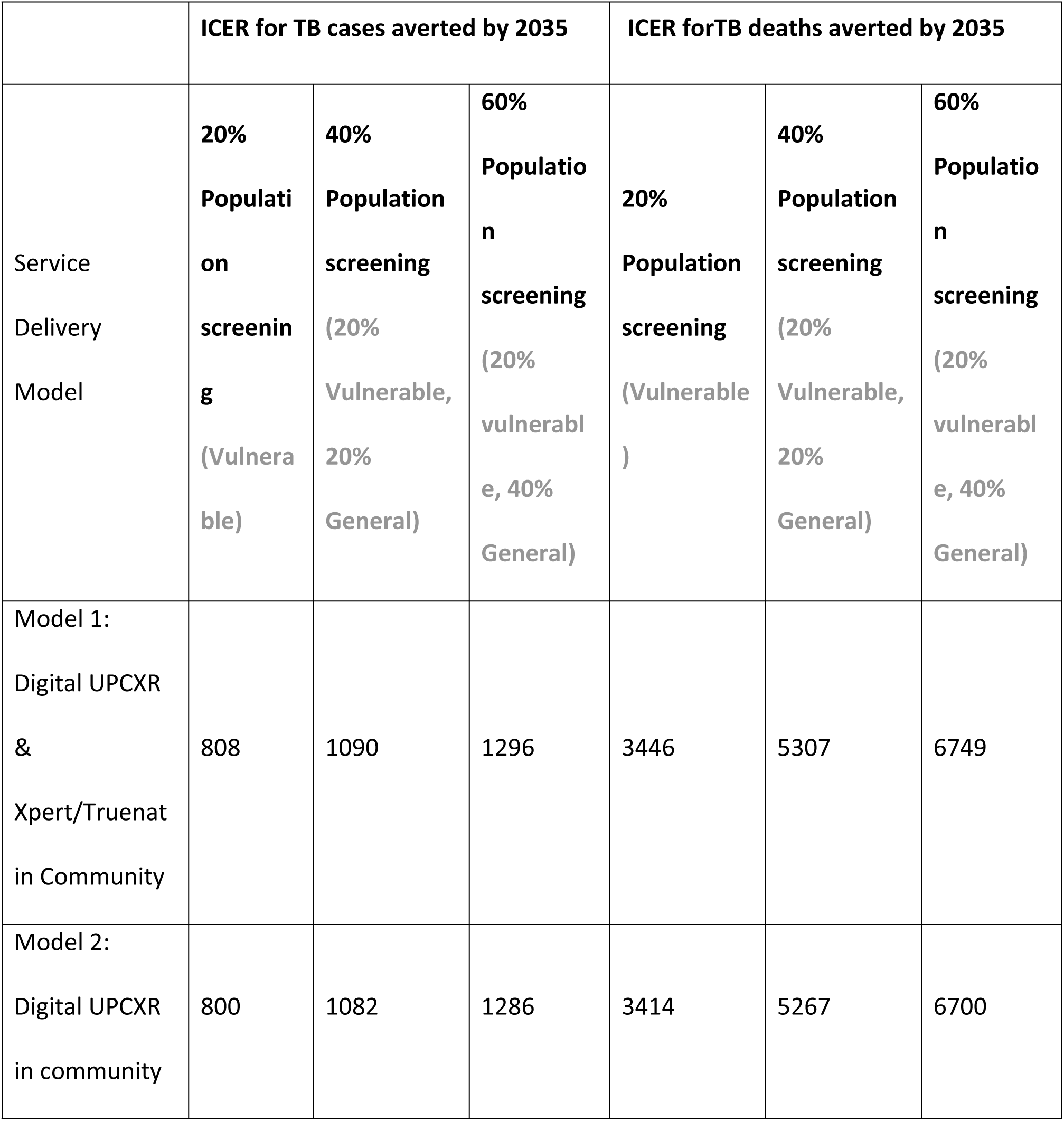
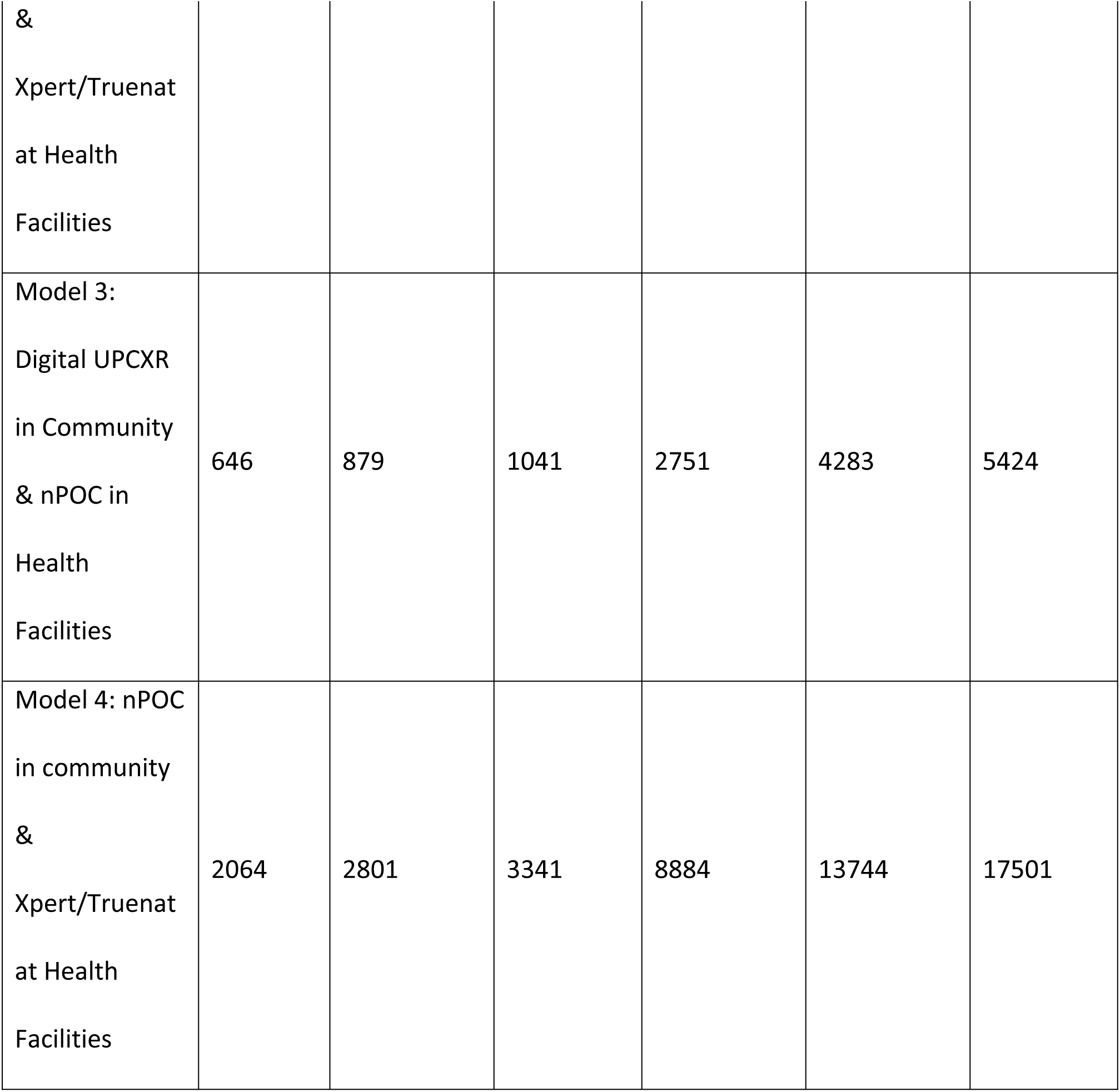
ICER for community-based TB screening scale-up with additional investment.

ICER per TB case averted was lowest at US$646 under Model 3 (digital UPCXR) in the community and nPOC testing at health facilities) at 20% population coverage (vulnerable population), and highest under Model 4 (nPOC testing in the community combined with Xpert/Truenat testing at health facilities) at 60% population coverage. The ICER per TB death averted followed a similar pattern to that observed for TB cases averted, with the lowest ratio again associated with Model 3 at the lowest coverage level and the highest with Model 4 at the highest coverage level. In terms of absolute impact, however, the largest number of TB cases and deaths averted were projected under Model 1 – digital UP-CXR combined with Xpert/Truenat testing in the community, at 60% population coverage.

## Discussion

This modelling analysis demonstrates that substantial reductions in TB incidence and mortality can be achieved across Asia’s ten high-burden countries through a combination of health facility strengthening and scaled-up population screening using existing and newly available diagnostic tools. However, the findings also underscore a critical message: even substantial expansion of current interventions is unlikely to be sufficient to meet the WHO End TB incidence targets for 2035 without the introduction of effective vaccines and additional preventive strategies [1].

Across all countries studied, strengthening of health facilities alone yielded meaningful declines in TB incidence and deaths, reflecting the importance of improved diagnostic coverage, reduced loss to follow-up, higher treatment success, and expanded use of TPT. These gains are consistent with previous modelling and empirical studies showing that closing gaps in the cascade of care can substantially reduce TB mortality in the short to medium term. Nevertheless, the projected impact on transmission was more modest, particularly in countries with large pools of undiagnosed or subclinical disease, highlighting the inherent limitations of predominantly passive case-finding approaches.

The addition of systematic screening of high-risk populations produced markedly larger projected reductions in both incidence and mortality. This finding reinforces growing evidence that TB burden is highly concentrated within vulnerable subpopulations and that targeted active case-finding represents an efficient and high-impact strategy [4,25–27]. By explicitly incorporating risk groups with higher progression and reactivation rates, the model captures the disproportionate epidemiological benefit of reaching these populations earlier in the disease course. Importantly, screening strategies incorporating UPCXR and Xpert/Truenat testing identified individuals who would otherwise remain undetected under symptom-based screening alone, thereby interrupting transmission chains and reducing future disease burden. Beyond the epidemiological gains, prioritising these populations carries an intrinsic equity rationale: the groups most affected by TB — people living in poverty and undernourishment, those with HIV, residents of overcrowded settlements, and displaced populations — are precisely those with the least access to passive case-detection services. Active screening strategies that included UPCXR and Xpert/Truenat testing to reach these communities therefore simultaneously advance both efficiency and health equity objectives. Expanding screening beyond high-risk groups to include segments of the general population generated additional, albeit diminishing, marginal returns. While screening up to 60% of the population resulted in further projected reductions in incidence and mortality, the incremental benefit relative to targeted screening was smaller, particularly in lower-incidence settings. However, the model shows that without screening of population beyond the high risk groups the End TB targets will not be reached. This finding has important programmatic implications: it suggests that prioritising vulnerable and high-transmission groups may offer a more efficient pathway to rapid impact, especially in resource-constrained environments. Decisions regarding the pace and sequencing of the broader population screening should therefore be guided by local epidemiology, operational feasibility, and cost-effectiveness considerations. Nevertheless, achieving End TB targets will ultimately require countries to progress beyond targeted approaches-broader population screening is not an alternative to efficient prioritisation but its natural progression as programmes mature and transmission is driven down.

Despite these substantial projected gains, none of the non-vaccine scenarios achieved the End TB incidence targets by 2035. This aligns with a growing consensus in the TB modelling literature that current tools, even when optimally deployed, are unlikely to end TB in high-burden settings over the next 10 years [6]. Encouragingly, several vaccine candidates — including M72/AS01E, which has demonstrated approximately 50% efficacy against active TB disease in Phase 2b trials — are advancing through late-stage development, making the vaccine scenarios modelled here a plausible near-term prospect rather than a distant aspiration. The introduction of a prevention-of-disease vaccine with moderate efficacy led to a step change in projected impact, pushing projected incidence reductions to approximately 80% relative to 2015 across all countries. Furthermore, the hypothetical addition of a prevention-of-relapse effect was sufficient to achieve WHO End TB targets in the model, underscoring the potential transformative role of vaccines that act at multiple points in the TB disease pathway.

The multi-country nature of this analysis highlights both shared patterns and important heterogeneity across settings. Countries such as India, Bangladesh, and Indonesia account for the largest absolute numbers of cases and deaths averted, reflecting their population size and disease burden, whereas smaller countries such as Mongolia and Tajikistan experience proportionally larger relative gains. These differences emphasise the need for country-specific prioritisation and tailored implementation strategies, even within a common regional framework.

The model indicated that the greatest epidemiological impact is achievable with AI-enabled digital UPCXR–based screening with Xpert/Truenat testing at the community level (Model 1) while digital UPCXR in the community combined with nPOC testing at health facilities (Model 3) represents the least-cost option. Although the nPOC in community & Xpert/Truenat at Health Facilities currently incurs the greatest cost, however in reality, this may change over time due to volume-based decreases in unit costs of nPOC tests, potentially altering the relative cost effectiveness of the service delivery options.

The cost-effectiveness findings warrant interpretation beyond relative model comparisons. The lowest ICER observed — US$646 per TB case averted under Model 3 at 20% vulnerable population coverage — compares favourably with estimates from comparable community-based TB screening programmes in high-burden settings, suggesting that targeted screening using available technologies represents good value for money even in the absence of formal QALY-based threshold analysis. Importantly, the ICER increases with each incremental expansion of coverage beyond high-risk groups, reinforcing the programmatic case for prioritising vulnerable populations as the most cost-efficient entry point. Countries seeking to maximise impact per dollar invested should therefore consider phased implementation — beginning with targeted vulnerable population screening before expanding to broader general population coverage as financing and operational capacity allow.

Several limitations should be considered when interpreting these findings. First, the model relies on assumptions regarding the size of vulnerable populations, relative risks, and screening performance that may vary within and between countries. Although uncertainty was explicitly incorporated through Bayesian calibration, data gaps, particularly for subclinical TB prevalence and private-sector care pathways, remain an important source of residual uncertainty. Second, the analysis assumes rapid and sustained scale-up of interventions with high coverage and quality, which may be challenging to achieve in practice due to health system constraints, workforce limitations, and social barriers to access. Third, vaccine scenarios are necessarily speculative, as product characteristics, rollout timelines, and real-world effectiveness remain uncertain. Fourth, the countries included in the analysis vary considerably in population, health care services and existing coverage and access. Applying a single national model with standardised costing assumptions necessarily simplifies this within-country heterogeneity; however, this approach was adopted to enable meaningful comparability of results across countries, and findings should be interpreted with this constraint in mind. Fifth, cost-effectiveness in this analysis is expressed as cost per TB case averted and cost per TB death averted rather than cost per QALY gained or DALY averted, which precludes direct comparison with standard willingness-to-pay thresholds. Within-study comparisons across service delivery models and coverage scenarios remain valid; however, future analyses incorporating age-stratified outcomes and disability weights would enable more direct comparison with accepted cost-effectiveness benchmarks.

Despite these limitations, this study provides a comprehensive and policy-relevant assessment of how combinations of existing and newly available tools could reshape TB epidemics across Asia’s high-burden countries. By integrating epidemiological impact with projections of cases and deaths averted across 10 high-burden countries, the analysis offers a robust evidence base to inform strategic planning and investment decisions. The results strongly support a shift toward proactive, technology-enabled screening strategies targeted at high-risk populations, alongside continued investments in health system strengthening and accelerated development of effective TB vaccines.

## Conclusions

Ending TB in Asia will require a decisive shift beyond incremental enhancements of existing programmes. This analysis demonstrates that large-scale, systematic screening, when coupled with timely and accurate diagnostics, can substantially accelerate declines in TB incidence and mortality, averting millions of cases and deaths over the coming decade. Targeted screening of vulnerable and high-risk populations emerges as the most efficient near-term strategy, offering greater cost-effectiveness than untargeted mass approaches while still generating substantial epidemiological impact. However, screening beyond the high-risk population is ultimately needed to reach the End TB targets. Health system strengthening alone, while meaningful, is insufficient to interrupt transmission at the scale required — community-wide active case-finding is an essential complement.

Among the service delivery models evaluated, AI-enabled digital UPCXR-based screening combined with Xpert/Truenat testing at the community level (Model 1) demonstrated the greatest epidemiological impact potential, while digital UPCXR in the community combined with nPOC testing at health facilities (Model 3) represents the least-cost option. An investment of USD 12.7 billion over the next five years in community-level implementation of digital UPCXR and molecular diagnostics could avert an additional 9.8 million TB cases and 1.9 million deaths across ten Asian countries over the following decade.

Despite these projected gains, none of the screening and health system scenarios modelled were sufficient to achieve WHO End TB targets by 2035. Reaching elimination will ultimately require integrating screening and health system investments with transformative preventive innovations — most critically, the development and large-scale deployment of effective TB vaccines. The hypothetical vaccine scenarios modelled here suggest that a prevention-of-disease vaccine with moderate efficacy could push incidence reductions toward 80% relative to 2015, and that the addition of a prevention-of-relapse effect could bring End TB targets within reach. Strategic investments made now across large scale screening, health system strengthening, and vaccine development have the potential to fundamentally reshape the TB epidemic trajectory across Asia.

## Data Availability

No data was generated by this study. The following existing data sources were used: [Epidemiological data] from [The Global TB Report] available via [https://www.who.int/teams/global-programme-on-tuberculosis-and-lung-health/tb-reports/global-tuberculosis-report-2025].

## Acknowledgements

This study was conducted as part of the *Ending Complex and Challenging Infectious and Tropical Diseases* (ExCITD) initiative, supported by the Asian Development Bank (ADB) under technical assistance (TA) 10728-REG. The findings, interpretations, and conclusions expressed in this publication are those of the authors and do not necessarily reflect the views of the Asian Development Bank or Stop TB Board.

## Author contributions

Conceptualization: Amandeep Singh, Suvanand Sahu

Investigation: Sandip Mandal, Kirankumar Rade

Methodology: Sandip Mandal, Kirankumar Rade, Sreenivas A. Nair

Project administration: Amandeep Singh, Sreenivas A. Nair

Writing – original draft: Sandip Mandal, Kiran Kumar Rade

Writing – review & editing: Amandeep Singh, Sreenivas A. Nair, Suvanand Sahu

## Supporting information captions

1. **Supplementary information: technical details**

## Notes

### Competing Interest Statement

The authors have declared no competing interest.

### Funding Statement

The author(s) received no specific funding for this work.

### Author Declarations

This study used publicly available, de-identified data and did not involve human participants directly. Therefore, ethical approval and informed consent were not required.

